# Comparing the impact on COVID-19 mortality of self-imposed behavior change and of government regulations across 13 countries

**DOI:** 10.1101/2020.08.02.20166793

**Authors:** Julian C. Jamison, Donald Bundy, Dean T. Jamison, Jacob Spitz, Stéphane Verguet

## Abstract

**Background:** Countries have adopted different approaches, at different times, to reduce the transmission of coronavirus disease 2019 (COVID-19). Cross-country comparison could indicate the relative efficacy of these approaches. We assess various non-pharmaceutical interventions (NPIs) over time, comparing the effects of self-imposed (i.e. voluntary) behavior change and of changes enforced via official regulations, by statistically examining their impacts on subsequent death rates in 13 European countries.

**Methods and findings:** We examine two types of NPI: the introduction of government-enforced closure policies over time; and self-imposed alteration of individual behaviors in response to awareness of the epidemic, in the period prior to regulations. Our proxy for the latter is Google mobility data, which captures voluntary behavior change when disease salience is sufficiently high. The primary outcome variable is the rate of change in COVID-19 fatalities per day, 16-20 days after interventions take place. Linear multivariate regression analysis is used to evaluate impacts. Voluntarily reduced mobility, occurring prior to government policies, decreases the percent change in deaths per day by 9.2 percentage points (95% CI 4.5-14.0 pp). Government closure policies decrease the percent change in deaths per day by 14.0 percentage points (95% CI 10.8-17.2 pp). Disaggregating government policies, the most beneficial are intercity travel restrictions, cancelling public events, and closing non-essential workplaces. Other sub-components, such as closing schools and imposing stay-at-home rules, show smaller and statistically insignificant impacts.

**Conclusions:** This study shows that NPIs have substantially reduced fatalities arising from COVID-19. Importantly, the effect of voluntary behavior change is of the same order of magnitude as government-mandated regulations. These findings, including the substantial variation across dimensions of closure, have implications for the phased withdrawal of government policies as the epidemic recedes, and for the possible reimposition of regulations if a second wave occurs, especially given the substantial economic and human welfare consequences of maintaining lockdowns.

## Introduction

Over the course of six months only, the transmission of the coronavirus disease 2019 (COVID-19) has spread to 188 countries worldwide: so far, COVID-19 has infected tens of millions of individuals and killed hundreds of thousands^1^. In the absence of available effective biomedical interventions like vaccines and treatments, and in anticipation of an unprecedented surge of patients in need of intensive care in hospitals, a large number of national responses have focused on the implementation of drastic non-pharmaceutical interventions (NPIs), including the closing of schools and universities, the prohibition of most commercial business, and the legal enforcement of local lockdowns and ‘shelter-in-place’ orders. As a result, in June 2020 an estimated 1.2 billion children who should have been attending schools were not doing so^2^, with long-term consequences for learning potential and the creation of national capital, and hundreds of millions of adults have had to cease their economic activities, with profound and immediate consequences for national economies and personal livelihoods and well-being. This is much more than a global health crisis^3^.

Now that the epidemic shows signs of receding in Western Europe, countries are urgently re-considering their NPI policies, especially when and how to suspend or reverse the school closure and ‘shelter-in-place’ policies that have such substantial developmental and economic consequences. The challenge, however, is that the method used to select the NPIs may be less helpful in planning their reversal. In the absence of real data or prior experience, the evidence base supporting the rollout of such unprecedented NPIs relied on mathematical forecasting models^4-7^ drawing on input parameters for epidemiologic quantities like severity and attack rate, risk factors, and timing of transmission, for which empirical validation remains nascent^8^. These assumptions may have been inadvertently misleading, hence need careful reassessment before being used as the basis for future decisions. For instance, with respect to school closures, a recent review^9^ and preliminary findings from France^10^ suggest that school children are not important drivers of coronavirus epidemics, in contrast to influenza, and school closure might play a substantially smaller role than the models had projected.

The need now is to retrospectively assess the true impact of NPIs on COVID-related morbidity and mortality, and to plan their withdrawal using empirical evidence. However, little work to date has conducted retrospective analyses of the possible mitigating effects of NPIs on the COVID death toll^11-16^. In this paper, we use an up-to-date time series of COVID-related mortality data from 13 comparable Western European countries. We undertake a statistical examination of the timing of introduction of NPIs and their impact on daily COVID deaths. Crucially, we include not only the full spectrum of government-mandated regulations but also measures of voluntary behavior change before the introduction of the government policies. This allows us to directly compare the effects of naturally-salient social distancing and enhanced hygiene practices, versus externally imposed and enforced regulations, with a view to contributing to the ongoing debate regarding re-evaluating school and workplace closures and other dimensions of reopening in Europe and beyond.

## Methods

### Data

Daily figures for new confirmed COVID-19 deaths by country were accessed through the European Centre for Disease Prevention and Control^17^. We used data for the 13 Western European countries with greater than 500 COVID deaths as of 16 May (Table 1), all of which had seven to eleven weeks of data, starting with t0 which is defined when the five-day moving average of daily deaths is first equal to at least five. This period captures all of the closure policies but (for simplicity) none of the subsequent relaxation of guidelines – where government and voluntary impacts are more difficult to disentangle.

**Table 1:** Summary statistics on COVID-related deaths and timing of measures of voluntary behavior and government-mandated regulations, for 13 Western European countries, March-May 2020

| Country | Total number of deaths | Total number of deaths per million population | Date of $t_0$ | Date mobility falls | Date mobility fall impacts deaths | Date national stay-at-home restrictions announced | Date national stay-at-home restrictions impact deaths | Number of days of data |
| --- | --- | --- | --- | --- | --- | --- | --- | --- |
| Austria | 628 | 71 | 24 March | 11 March | NA | 6 March | 22-26 March | 56 |
| Belgium | 8959 | 784 | 19 March | 12 March | 28 March – 1 April | 18 March | 3-7 April | 61 |
| Denmark | 537 | 93 | 23 March | 12 March | 28 March – 1 April | 13 March | 29 March – 2 April | 57 |
| France | 27529 | 411 | 08 March | 12 March | 28 March – 1 April | 17 March | 2-6 April | 72 |
| Germany | 7881 | 95 | 18 March | 9 March | NA | 9 March | 25-29 March | 62 |
| Ireland | 1518 | 313 | 27 March | 11 March | 27-31 March | 26 March | 11-15 April | 53 |
| Italy | 31610 | 523 | 01 March | 24 February | 11-15 March | 10 March | 26-30 March | 79 |
| Netherlands | 5643 | 327 | 16 March | 10 March | 26-30 March | 12 March | 28 March – 1 April | 64 |
| Portugal | 1190 | 116 | 23 March | 12 March | 28 March – 1 April | 19 March | 4-8 April | 57 |
| Spain | 27563 | 590 | 08 March | 11 March | 27-31 March | 14 March | 30 March – 3 April | 72 |
| Sweden | 3646 | 358 | 23 March | 11 March | 27-31 March | NA | NA | 57 |
| Switzerland | 1594 | 187 | 19 March | 10 March | 26-30 March | 17 March | 2-6 April | 61 |
| United Kingdom | 33998 | 511 | 14 March | 12 March | 28 March – 1 April | 23 March | 8-12 April | 66 |

Notes: Deaths data cumulative up to and including 16 May. We describe policy changes as taking effect over a 5-day date range because our model uses a 5-day rolling average. $t_0$ denotes the date at which the 5-day rolling average reached five deaths, which we define as the start of the epidemic.

COVID mortality data were used because: death constitutes a significant event; death certifications are less likely (than case notifications) to suffer from misclassifications; and the completeness of death data is far greater than that of case notification data due to varying testing capacity and accuracy across countries. However, (i) actual death tolls are still likely to differ from currently reported figures due to reporting issues; (ii) recording protocols can affect total numbers (for example whether deaths in nursing care homes are included); and (iii) reported date of death can be delayed from the actual date of death. Issues (i) and (ii) are mitigated here by focusing on relative changes in deaths, which also allows us to abstract away from total population size. Issue is mitigated in part by taking a five-day moving average of deaths. Hence, as our dependent variable, we study the evolution over time of the following percentage change in smoothed daily deaths:

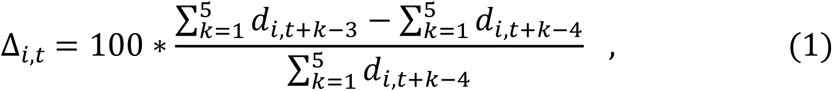

where d_*i,t*_ is the daily reported number of deaths in country *i* on day *t*.

To get a sense for the behavior of this variable ∆_*i,t*_, note that early in the pandemic the number of deaths per day is typically rising, corresponding to the number of new infections having been growing a few weeks earlier, which implies that ∆_*i,t*_> 0. Late in the pandemic, when the number of daily deaths is declining, this percent change will be negative. In between, each day will yield approximately the same number of deaths and hence our dependent variable will be around zero. Smaller values are always better, since they imply a slower rise in fatalities (if positive) or a more rapid decline (if negative). Table A1 in the supplementary appendix shows the distribution of values of this variable in our data, week by week.

For the interventions, we focus on two broad categories: government-imposed policies and regulations, versus self-imposed and voluntary actions. The Oxford COVID-19 Government Response Tracker provides dates and intensities for multiple categories of government policies across the globe^18^. Here, we focus on their ‘containment and closure’ categories: school closing; workplace closing; canceling public events; restricting public events and gathering sizes; closing public transport; stay-at-home (or ‘shelter-in-place’) requirements; and restrictions on internal movement and international travel. We define two alternate versions of the government *closure* measure: first, an easy-to-interpret *binary* one, which is directly comparable to the voluntary measure defined below, occurs when broad stay-at-home restrictions are initially promulgated; and second, a *continuous* closure measure which is simply the sum of scores across all categories, normalized by dividing by the maximum such score in the database.

Importantly, we also look at self-imposed restrictions on behavior which arose prior to the introduction of governmental interventions. Our primary measure, *mobility decline*, is based on Google’s Community Mobility Reports^19^, which assess physical mobility along different dimensions, as compared to a pre-crisis baseline within each country. Our indicator (dummy) variable switches from 0 to 1 in a given country when the mobility index is negative (representing activity being below baseline levels) *and* remains so thereafter, for all of the three mobility categories corresponding to workplaces, transit stations, and retail and recreation. Figure 1 presents mobility data for three illustrative countries. We do not consider residential mobility, since that does not necessarily correspond to social distancing. Similar changes were observed in China early in the pandemic, where regional air pollution, indicative of reduced traffic and production, decreased after cases were reported locally but before any government restrictions had been imposed^20^.

**Figure 1:**
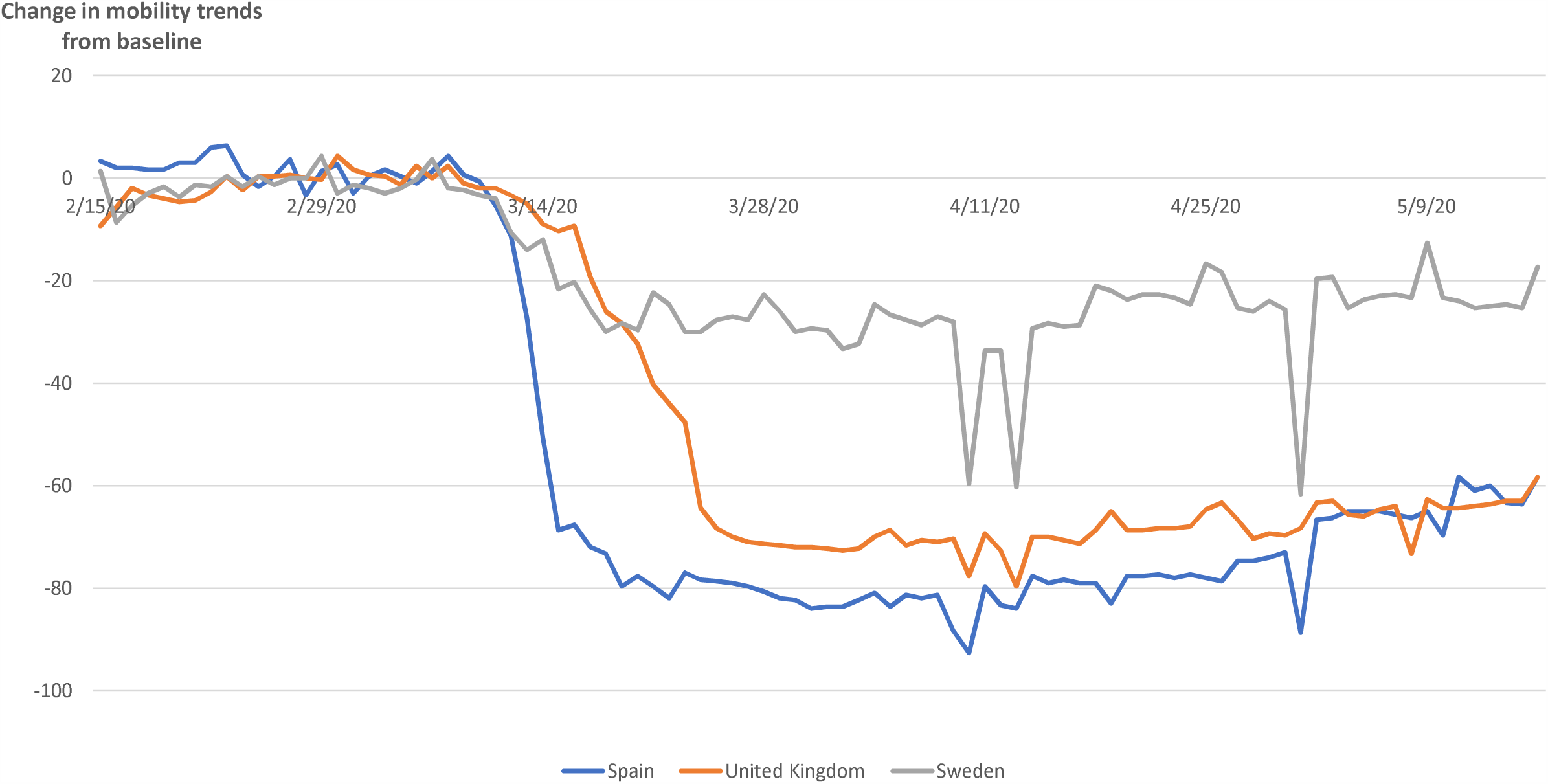
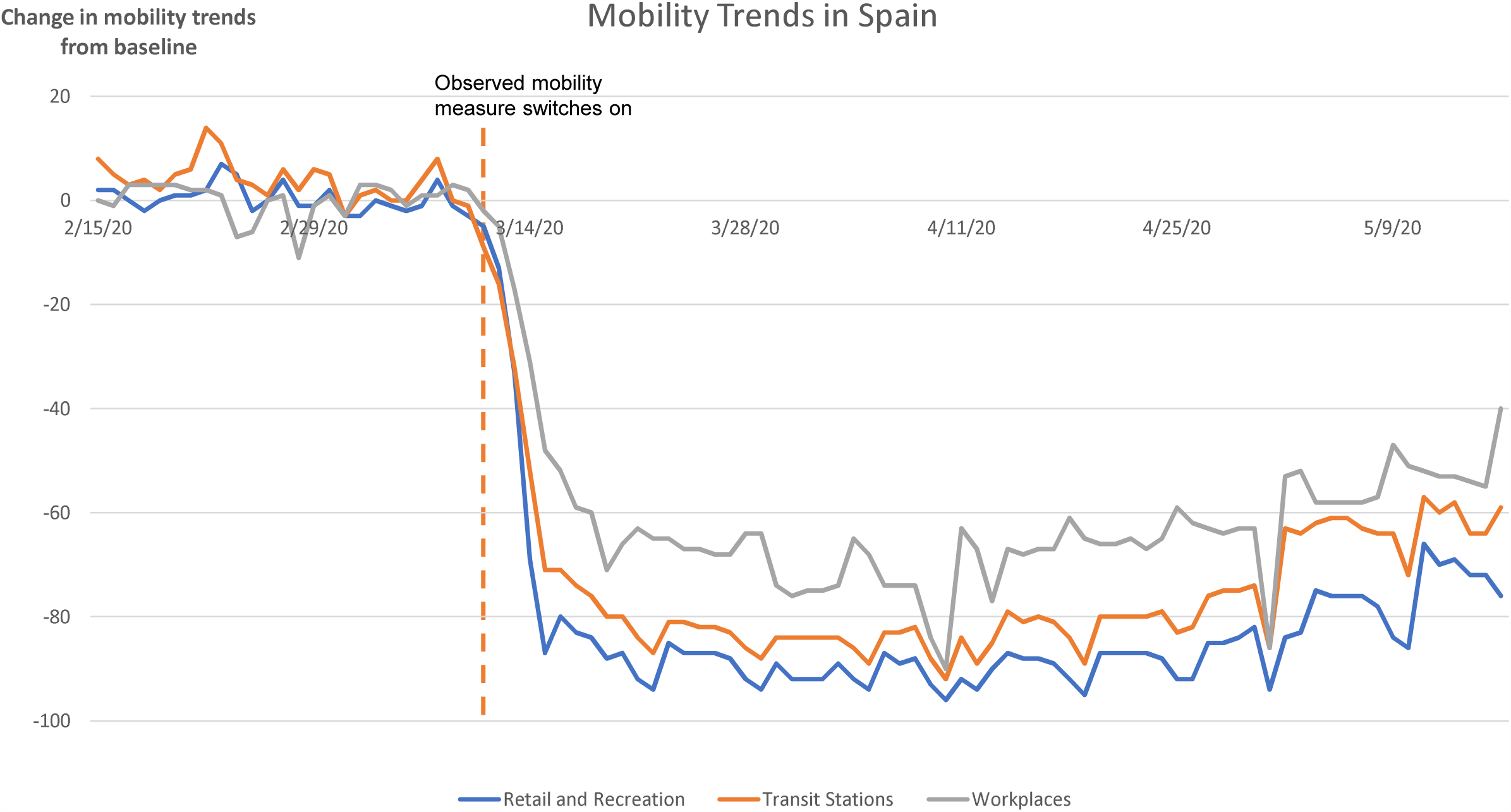
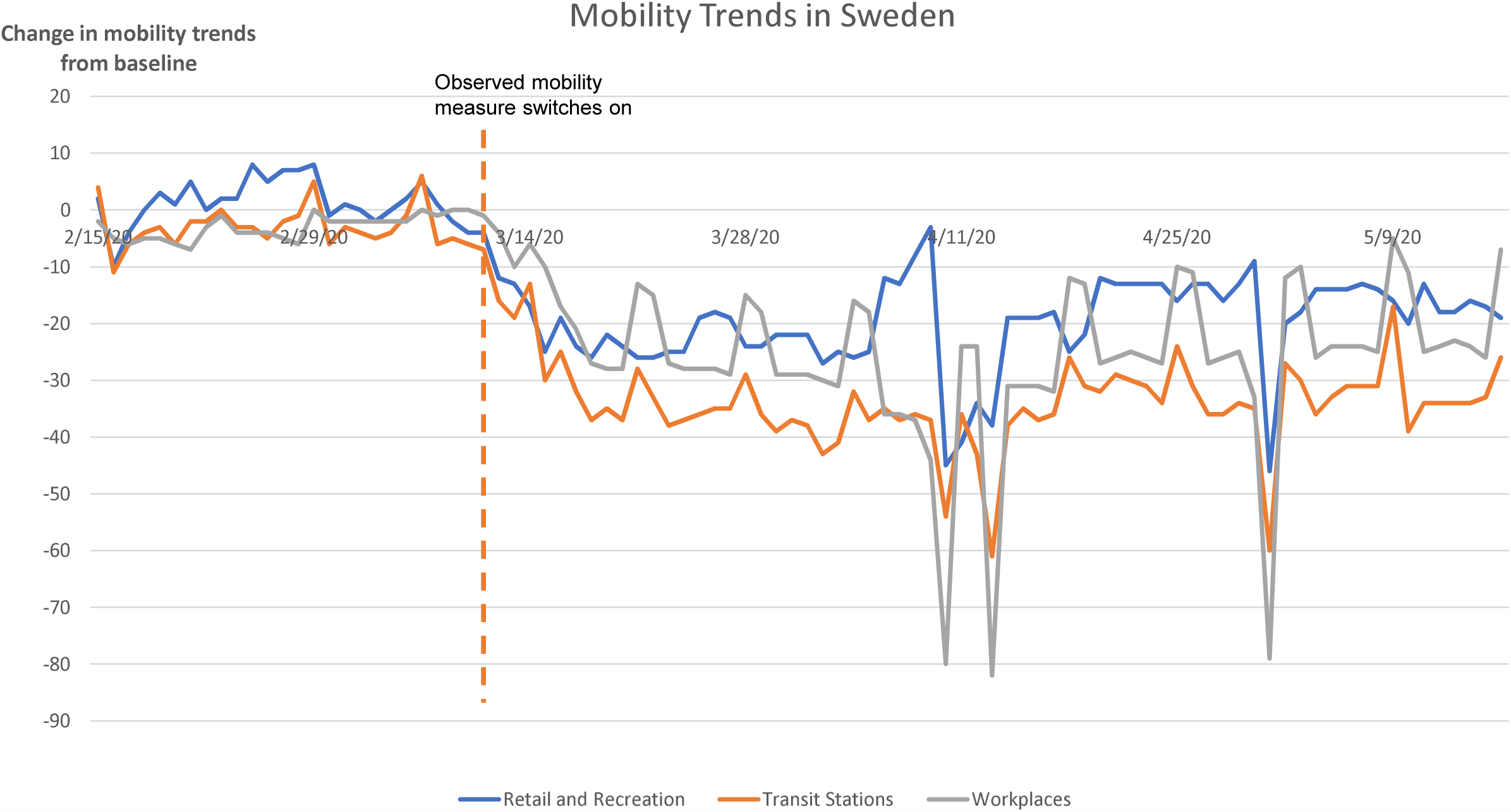
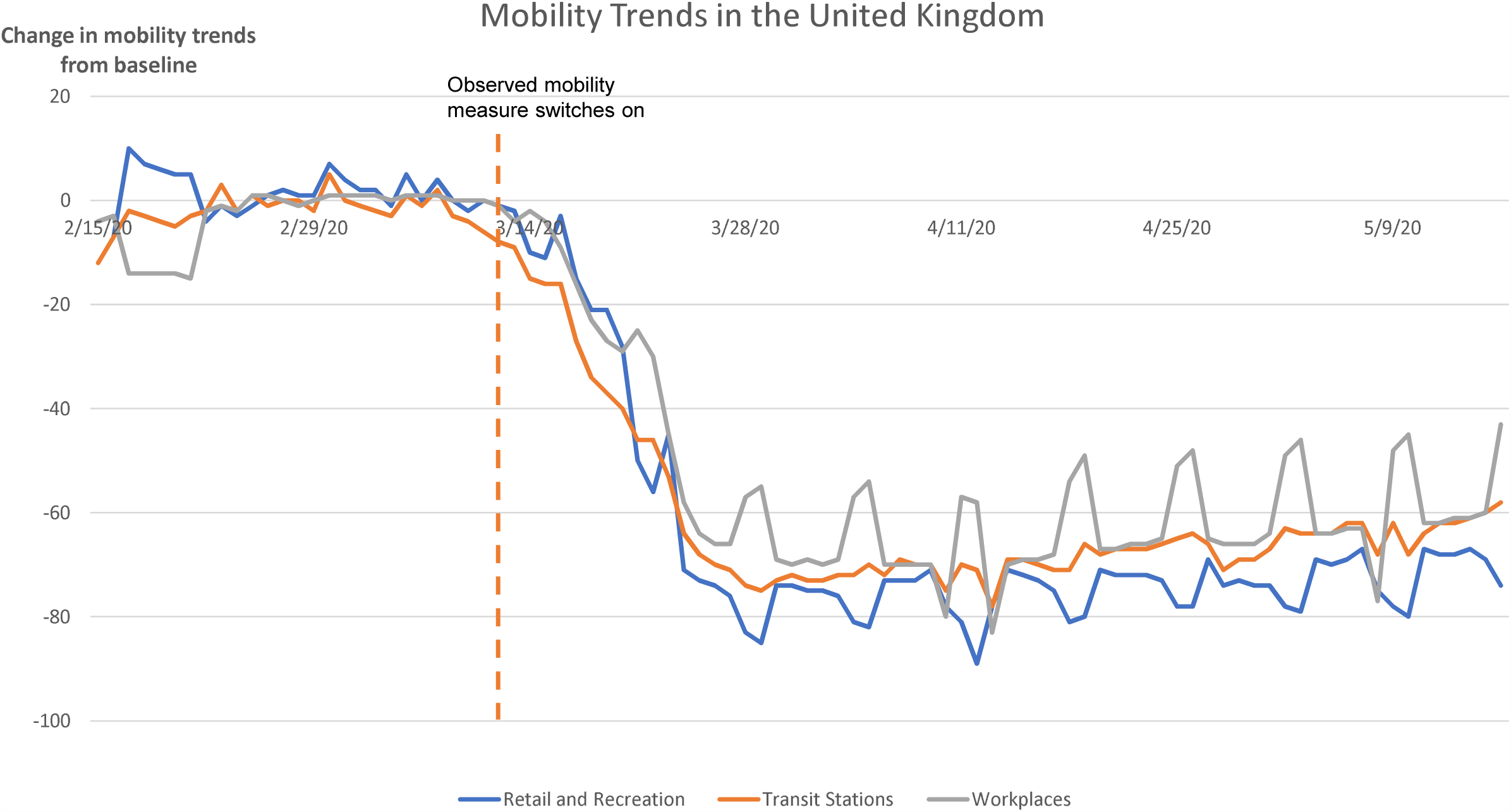
Mobility Trends in Spain, Sweden and the United Kingdom.

The mobility indicator switches back from 1 to 0 when the government binary closure indicator turns on in that country, because the goal is to evaluate the effect of naturally-occurring behavior change. If binary closure takes place before self-imposed mobility decline (as in Austria and Germany), then the mobility variable remains equal to 0 throughout the study period. As with closure, we also define a continuous version of this variable equal to the normalized sum of mobility decline across the three relevant categories, on a given date.

Evidently, none of these interventions, either regulatory or voluntary, will have an immediate effect on fatalities due to COVID-19. Rather, we hypothesize that they will change the rate of new infections, leading to a change in deaths some time later. In order to model that delay, we assume that it is the sum of the incubation period, estimated to be 5 days^4^, and the period from symptom onset to death (for those who die), which has an observed median of 13 days^21^. Note that the overall typical time to death will be different from 13 days, because in a growing epidemic proportionately more observations are from recent infections (some of whom will die later). We are modeling the observed data in the midst of a growing epidemic, hence it is precisely the raw data that we need to match. Thus we assume a median lag of 18 days from time of intervention to time of death. There is naturally some distribution for this lag, thus we employ a 5-day moving average of deaths, corresponding to lags from 16-20 days (Table 1).

Figure 2 shows the 5-day average deaths for three illustrative countries, along with the median dates at which binary closure and (if relevant) binary mobility decline are assumed to have taken effect. The three countries include Spain and the UK, which both have relatively large populations, but where the local epidemic started relatively early and relatively late respectively, as well as Sweden, which is unique in our sample in that the government never imposed stay-at-home regulations.

**Figure 2:**
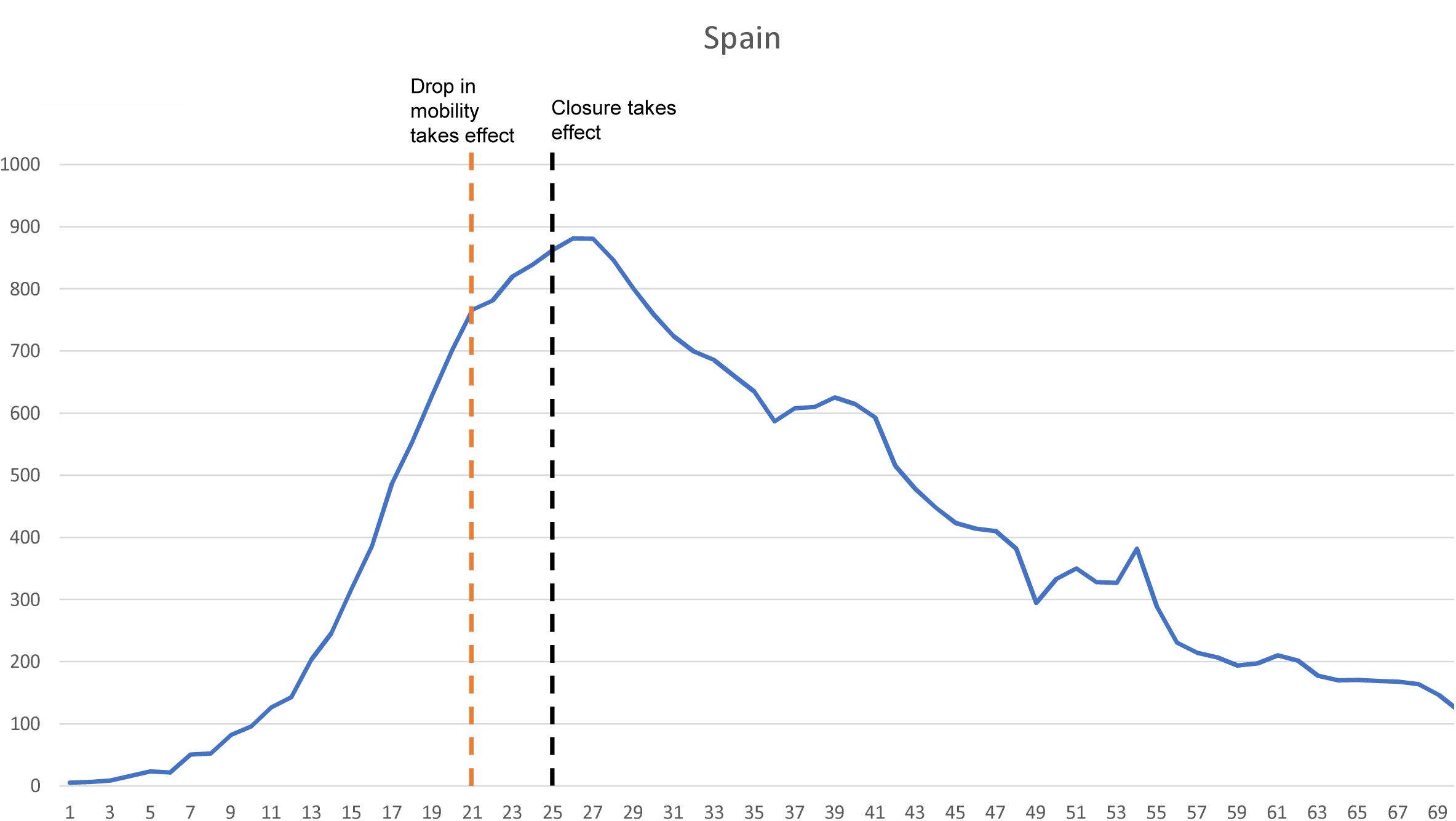
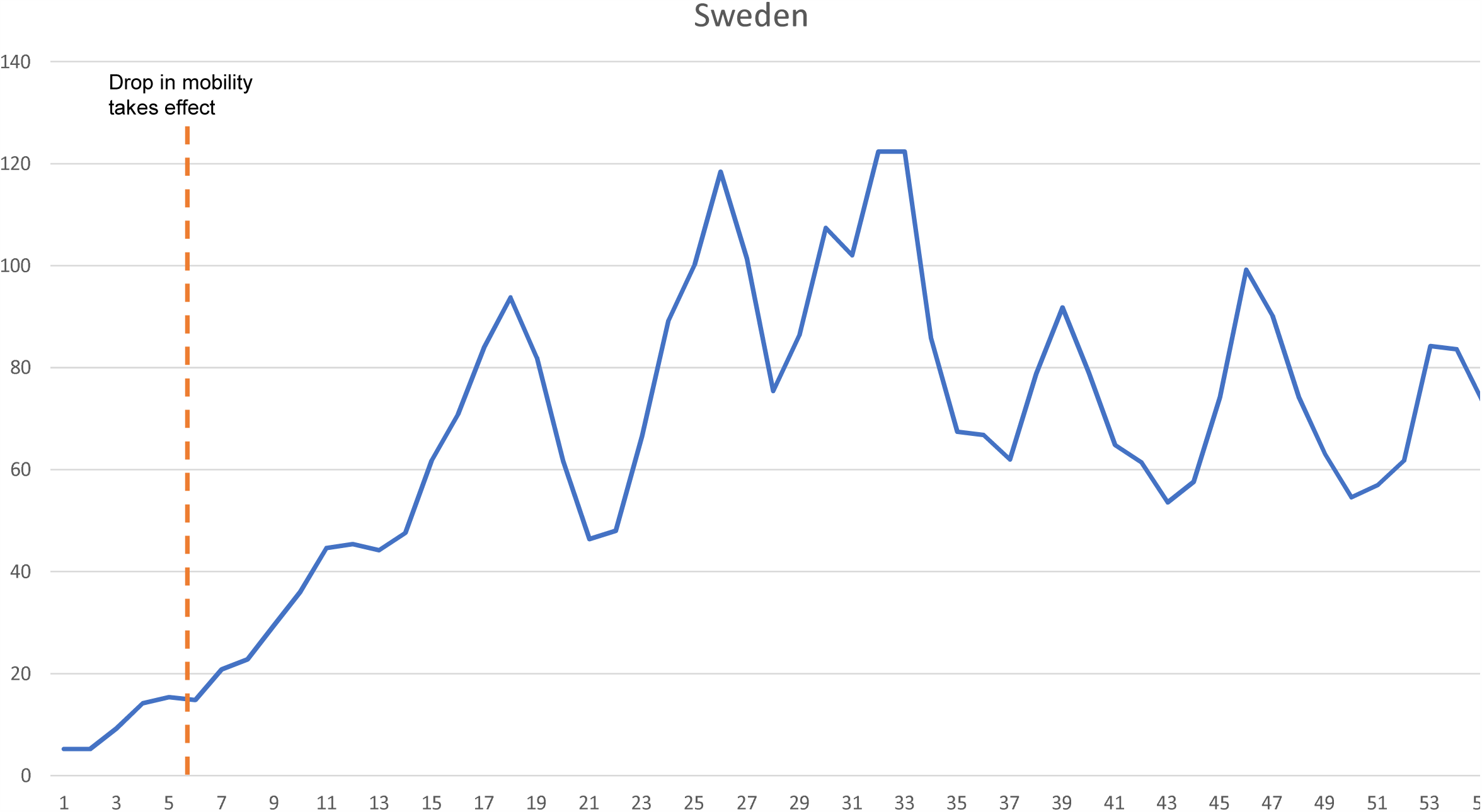
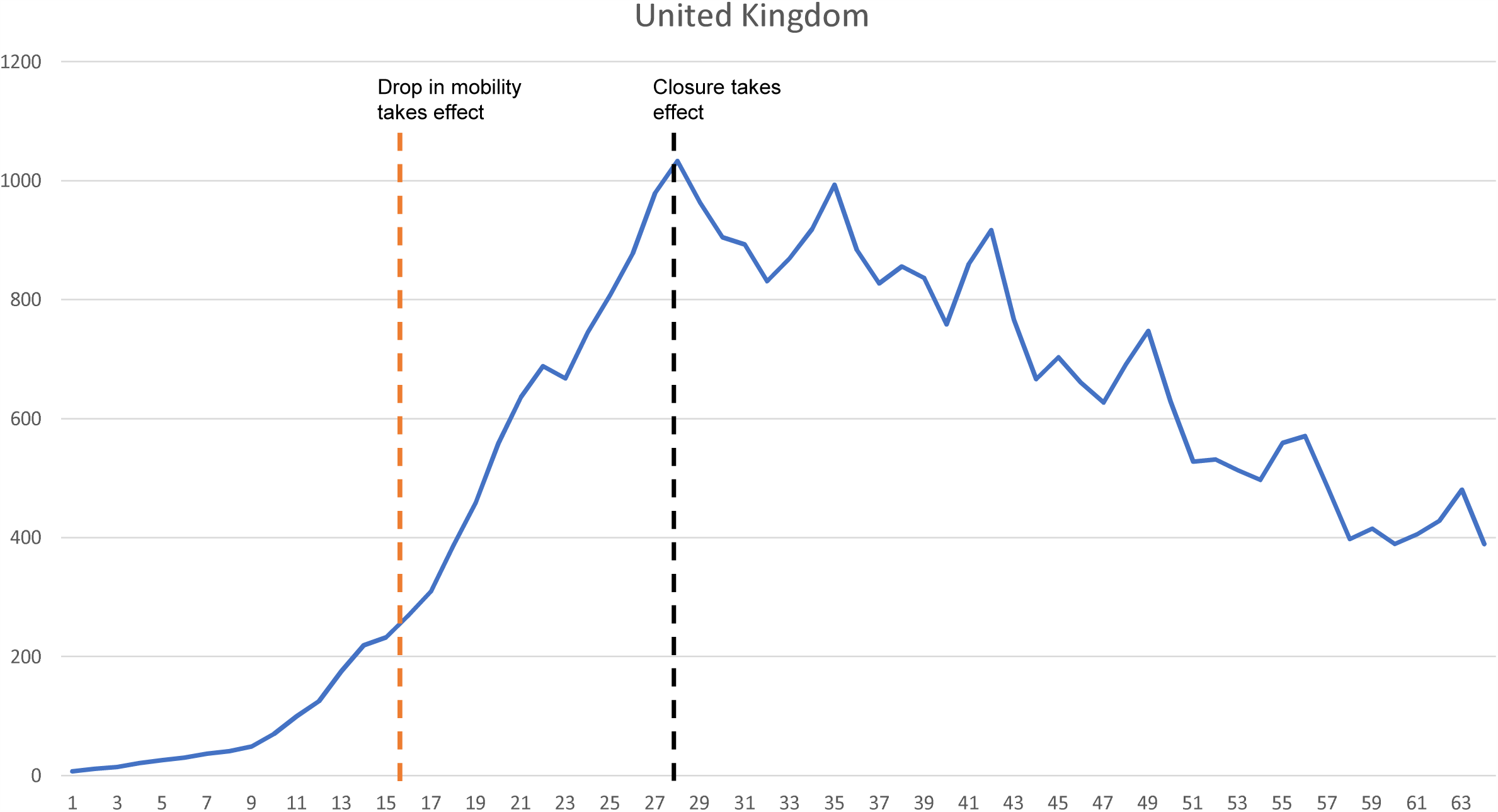

### Modeling approach

We evaluate the effect of NPIs on the rate of change in COVID deaths via a linear regression model. We study the evolution over time of the daily percentage change in deaths, using a random effect specification to further net out a range of country-specific factors that might affect both the total number of deaths and local behavioral and policy changes, such as the scale of the epidemic, political ideology, or infrastructure differences. We employed the following model specification for the daily percentage change in COVID deaths in country *i* at time *t*:

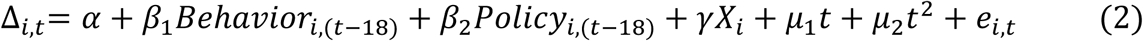

*Bhavior*_*i*,(*t*-18)_ and *Policy*_*i*,(*t*-18)_ are indicators of behavioral and policy changes, respectively: they are lagged by 18 days to reflect when policy changes materialize, as detailed above. *X*_*i*_ is a vector of country-specific controls: share of population over 65^22^, population density^22^, the number of acute care beds per 100,000 population^23^, as well as the starting date of the epidemic in each country (Table 1). Lastly, *t* counts the number of days since the start of the epidemic in each country: this captures exogenous time trends, as well as any endogenous learning by both individuals and clinicians. Because this effect is expected to be nonlinear, we also include *t*^2^ as an independent variable.

In order to help interpret the implications of the effect sizes, we estimated the number of days for deaths to double, as of one week into the epidemic, under the assumptions of no intervention; closure only; and salience only. To do this, we fixed *t* =7, predicted the expected country-average growth rate 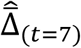 in each of the three conditions respectively, and applied the formula for doubling time: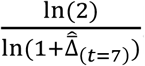

Finally, we estimated the number of COVID deaths that would have occurred in the first seven weeks from the local starting date, under analogous situations: no intervention; closure only (starting at *t*=7); and salience only (starting at *t*=7). This was calculated by summing deaths from *t*=0 (normalized to our threshold of 5) through *t*=50, after iterating forward using the modeled growth rates in each of the three hypothetical policy configurations. This is done at the level of individual countries, which cannot be directly compared to one another using our analysis but are illustrative of what would occur as a function of different underlying conditions. In particular, the primary cross-country variation in our model comes from the starting calendar date of the epidemic.

All statistical analyses used STATA/SE version 13.0. All data comes from publicly available aggregate sources, so no ethical approval was required. No external funding was utilized during the course of the study.

## Results

Table 2 presents the main results, with Model I being the preferred specification. The first row shows that as time passes, regardless of any external intervention, death rates go down. Specifically, for each day that passes, the daily change in fatalities goes down by 0.88 (0.67-1.10) percentage points. For similar reasons (results not shown), countries that experienced the start of the pandemic earlier -- such as Italy and Spain -- fare worse than those that experienced it later -- such as Germany and The Netherlands.

**Table 2:** Effect of Observed Mobility and Government Closure on Daily Change in Deaths, for 13 Western European countries, March-May 2020.

|  | Model I | Model II | Model III | Model IV |
| --- | --- | --- | --- | --- |
| Days from $t_0$ | -0.88 (0.000) | -0.86 (0.000) | -0.90 (0.000) | -0.81 (0.000) |
|  | [-1.1, -0.67] | [-1.1, -0.67] | [-1.1, -0.70] | [-1.02, -0.60] |
| Binary mobility | -9.2 (0.000) | -10.2 (0.000) | -9.0 (0.000) |  |
|  | [-14.0, -4.5] | [-14.7, -5.7] | [-12.7, -5.4] |  |
| Continuous mobility |  |  |  | -8.8 (0.022) |
|  |  |  |  | [-16.3, -1.3] |
| Binary closure | -14.0 (0.000) | -15.2 (0.000) | -13.1 (0.000) |  |
|  | [-17.2, -10.8] | [-18.7, -11.7] | [-15.5, -10.7] |  |
| Continuous closure |  |  |  | -18.7 (0.000) |
|  |  |  |  | [-23.1, -14.4] |
| Country fixed effects? | No | No | No | No |
| Number of observations | 778 | 778 | 778 | 776 |
| Notes: p-values are presented in parentheses, 95% confidence intervals are presented in square brackets. Specifications also included controls for $t$ squared, the percentage of population aged over 65, the population density, and number of acute care beds per 100,000 people. Standard errors are clustered at the country level. Observed mobility is the binary measure based off Google mobility data; continuous mobility is a measure calculated by summing the same three measures of the Google Mobility Index and normalizing across countries; binary closure is our binary variable based on the Oxford Policy Tracker index for stay-at-home restrictions; and continuous closure is a measure calculated by summing all 8 'containment and closure' categories in the Oxford Policy Tracker and normalizing across countries. We make one small change to the Oxford data: defining the German lockdown as being nationwide instead of regional. N is lower in Model IV because the lagged mobility data is only available for Italy from the third day of the epidemic. |  |  |  |  |

The central finding is that both the voluntary measure and the closure measure have substantial and highly statistically significant impacts on death rates 18 days later. A binary reduction in self-imposed mobility is associated with a 9.2 (4.5-14.0) percentage point reduction in the daily rate-of-change in fatalities, while binary government restrictions are associated with a 14.0 (10.8-17.2) percentage point reduction. This means that if deaths were growing by 5% per day, then voluntary behavior change would cause them to start declining by 4.2% per day, while government regulations would cause them to decline by 9% per day.

Models II and III estimate the same specification but with fatality lags of 17 and 19 days respectively, confirming that the results are robust across modeling choices. Model IV shows that the continuous measures also yield similar results. The closure measure in this case has a larger magnitude because it can be interpreted as all government policies being enacted at the maximal observed level simultaneously. The supplementary appendix reports further sensitivity analyses: first a specification of the main results with country fixed effects; second an alternate proxy for voluntary behavior change, occurring when the number of national deaths surpasses a salience threshold; and third the equalization of the length of data across countries.

Table 3 extends this analysis to separately examine the effect arising from different components of the continuous closure metric. Model I looks at all eight categories individually, finding that three of them are significant at the 5% level: closing non-essential workplaces is estimated to reduce the change in deaths by 4.7 (0.3-7.1) pp; restricting public events reduces it by 5.7 (1.5-9.9) pp; and limiting international travel reduces it by 4.8 (0.7-9.0) pp. Models II and III combine similar categories to examine consistency across domains, until finally model IV is the fully aggregated version replicating model IV in Table 2. This suggests that the results are stable across the combined versions, with the possible exception of limiting public events (which no longer appears effective when combined with restrictions on the size of gatherings).

**Table 3:**
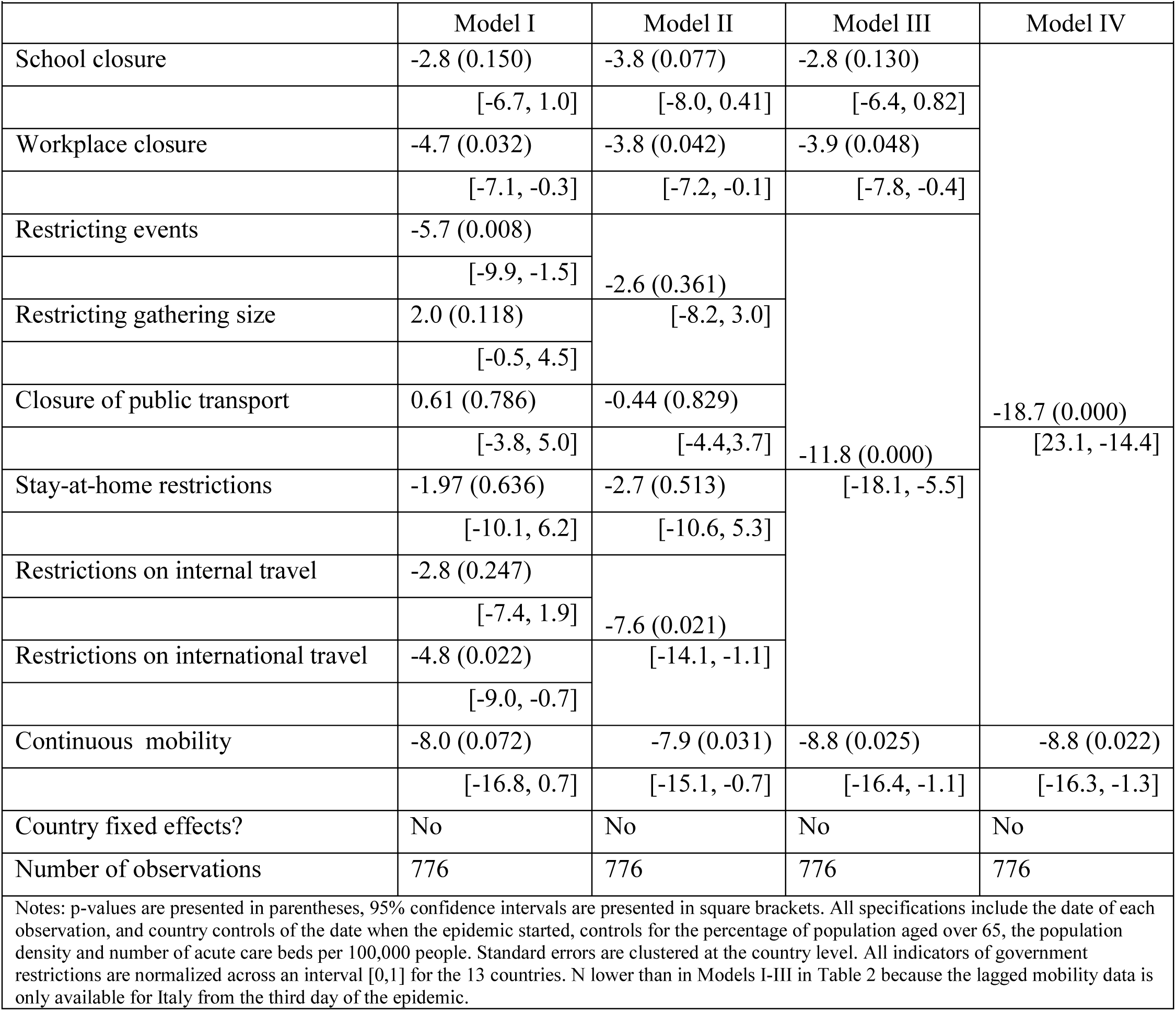
Disaggregated impact of the various NPIs on Daily Change in Deaths, for 13 Western European countries, March-May 2020.

In order to help interpret the magnitudes of the effect sizes, we use the coefficients in Model I of Table 2 to estimate the number of days for deaths to double, as of *t* = 7, under three scenarios: no intervention of any kind, self-imposed behavior change, and government-enforced closure. Doubling time is increased from 3.0 (2.7 – 3.4 days) to 4.5 days (3.7 – 5.8 days) with a voluntary reduction in mobility, or to 6.1 days (5.0 – 8.1 days) with government lockdown.

Finally, Table 4 compares our projections of the expected number of deaths under three different scenarios – no intervention, salience from *t* = 7, and closure from *t* = 7 – with reality for exemplar countries. Spain experienced an early local start to the pandemic and we estimate that it would have had 174,935 deaths in the first 50 days if there had been no interventions at all. In reality it had 23,467 deaths over that period – but if it had closed down two weeks earlier, deaths would have been only 3,487. Even with purely self-imposed changes throughout, if they had begun early, the number of deaths would have been only 11,430. Thus the timing of interventions is crucial, more important than the severity: tens of thousands of lives could have been saved even without a full lockdown. Meanwhile Sweden experienced a later start and hence is estimated, in the absence of any interventions, at 53,528 deaths in its first 50 days. Reality was 3,271 deaths – similar to our projection in the case of voluntary changes only (since that was indeed what happened there, although the timing was slightly different) – while government closure could have reduced this further to 1,494.

**Table 4:**
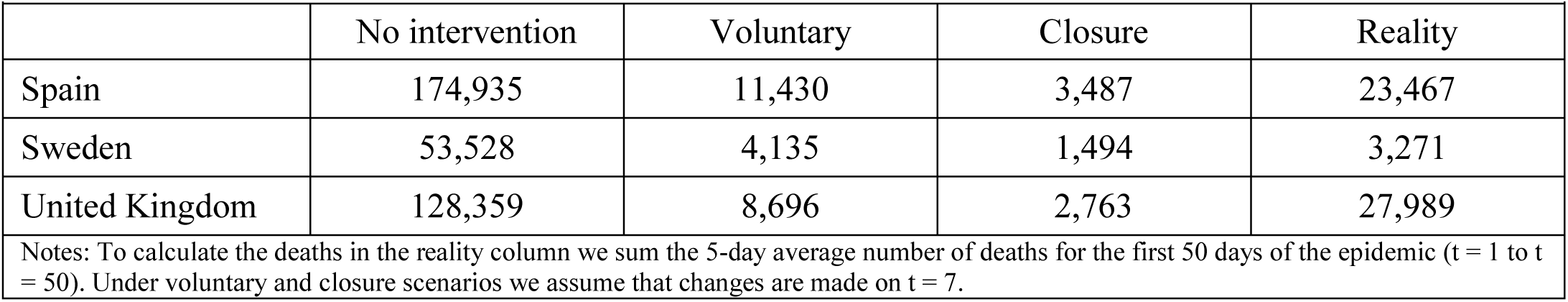
Predicted Number of Deaths During the First 50 days of the Epidemic Under Different Scenarios

|  | No intervention | Voluntary | Closure | Reality |
| --- | --- | --- | --- | --- |
| Spain | 174,935 | 11,430 | 3,487 | 23,467 |
| Sweden | 53,528 | 4,135 | 1,494 | 3,271 |
| United Kingdom | 128,359 | 8,696 | 2,763 | 27,989 |

Notes: To calculate the deaths in the reality column we sum the 5-day average number of deaths for the first 50 days of the epidemic ( $t = 1$ to $t = 50$ ). Under voluntary and closure scenarios we assume that changes are made on $t = 7$ .

## Discussion

Using daily death data from 13 European countries, we find that NPIs can substantially reduce fatalities from COVID-19. Both self-initiated behavior change prior to official government guidelines, as well as more strict regulations themselves, had a significant effect on the ensuing rate of change of deaths. Intriguingly, the magnitudes of the two approaches are not vastly different from one another: government closures reduce the growth rate of deaths by around 14 percentage points, while behavior change in the absence of government pronouncements reduces the growth rate by just over 9 percentage points. Either approach, applied sufficiently but realistically early, would have saved thousands of lives in a typical country in our sample.

Most previous observational analyses of NPIs – primarily stay-at-home and social distancing directive – have focused on cases rather than fatalities, either in individual countries^24-27^ or across multiple countries^28-29^. There is limited existing work regarding the effect specifically of stay-at-home policies on deaths, in Brazil^11^, France^12-13^, and the United States^14-15^, but not of the full range of government interventions. The most similar analysis to this paper comes from Imperial College^16^, which examines the impact of regulations on fatalities in 11 European countries. They assess multiple government interventions (including explicit encouragement of social distancing) but do not consider voluntary behavior change. Their main result is that lockdowns had a strong impact but that no other policies (social distancing, limiting public events, closing schools, or self-isolation) had a significant effect at all. Our conclusions are very different, perhaps due to the fact that we take a naïve but direct statistical approach to the relationships rather than filtering them through a complex structural model. One other paper^30^ examines the impact of multiple interventions on cases (not deaths) across six countries globally, using a similar reduced-form approach to that taken here. However like the others it does not consider voluntary changes, so its counterfactual scenario (exponential growth of 38% per day in the absence of policy) becomes less and less realistic over time, exaggerating the role of explicit policies in flattening the curve.

To our knowledge, no previous study has explored the remarkable impact of self-imposed behavior change, yet these empirical results suggest that enforced lockdown regulations offer only modestly stronger epidemiological outcomes than well-timed voluntary behavior change. This is an important observation in the context of addressing future spikes or epidemics in other countries, especially given that self-motivated behaviors – such as social distancing, improved personal hygiene, reducing unnecessary travel, and working remotely when possible – are intrinsically less disruptive and more individually malleable than regulatory options – such as shelter-in-place orders, closing schools, and banning public transit. Most importantly, as the pandemic is expanding in low- and middle-income countries in regions like South Asia and sub-Saharan Africa where the health systems are often weak, where the sociodemographic and cultural traits differ immensely, and where the great majority of people live on a daily wage, weighing the feasibility along with the full benefits and costs of NPIs will be critical^31^.

The results also provide insights into which of the government regulations were more effective when we disaggregate government policies into various subcategories. Limiting travel, particularly international travel, seems to have a significant effect, as does closure of non-essential workplaces. However other categories, including closing schools and imposing stay-at-home rules, show smaller and statistically insignificant effects. These subcategory findings are to be interpreted with caution, in part because there is less variation than optimal across countries in our sample, but they do suggest that policy-makers should think carefully about whether – and if so for how long – the various restrictions, some of which are known to inflict large costs on health, education, welfare and the economy, are necessary.

As one of the first studies to explore these issues empirically using substantial data, this analysis inevitably involves a number of limitations. Without randomization or other exogenous variation in the treatments, evidently, we cannot fully ascertain a causal link between the NPIs and the resulting changes in death rates. We do not expect any reverse causality, since future deaths will not change current behaviors. However, it is conceivable that some third variable, for instance heightened media attention and scrutiny, could directly influence both government policies and individual behaviors. Future studies, using data on this and similar potential confounders, may be able to fully disentangle the various mechanisms at play. Finally, the quality of the fatality data is subject to variation in reporting standards across countries, although this will be partially mitigated by focusing on rates of change rather than levels. Similarly, the quality of the government closure data is, although compiled independently and not biased in any way, somewhat subjective in nature as to the precise degree of severity in each category at each point in time.

The main messages here are that NPIs can have significant impacts in reducing COVID-19 mortality, and that the empirical evidence suggests that the simpler, more flexible and more easily reversed interventions that result in self-motivated voluntary behavioral change, working remotely to the extent possible, and reducing large-scale discretionary travel, are as effective as stricter officially imposed regulations. Precisely why that is we cannot say from this analysis, but the distinction may be important to those countries still in the earlier stages of responding to the pandemic, particularly in South Asia and sub-Saharan Africa. Effective planning and evidence-based policy is that much more important for countries where finances are limited, health systems are weak, and the majority of people rely on a daily wage.

## Data Availability

All data is publicly available and is linked to in the manuscript references.

## Contributors

JJ conceived the study, with input from the other authors. JS created the merged dataset and analyzed the data. JJ and SV wrote the first draft. All authors contributed to the final draft.

## Declaration of interests

All authors declare no competing interests.

## Supplementary appendix

**Table A1:** Percentile distribution of daily change in smoothed deaths, by week

|  |  |  |  |  |  |  |  |  |
| --- | --- | --- | --- | --- | --- | --- | --- | --- |
| Percentile | Overall | Week 1 | Week 2 | Week 3 | Week 4 | Week 5 | Week 6 | Week 7 |
| 10th | -15.9% | 0.0% | -1.8% | -7.6% | -9.6% | -16.5% | -17.8% | -22.7% |
| 25th | -6.1% | 9.6% | 7.7% | -1.2% | -4.0% | -6.2% | -13.6% | -13.0% |
| 50th | 1.5% | 21.6% | 15.4% | 8.1% | 0.4% | -2.0% | -4.6% | 5.2% |
| 75th | 12.8% | 39.5% | 24.3% | 13.6% | 8.5% | 4.5% | 2.5% | 5.3% |
| 90th | 25.4% | 57.1% | 32.3% | 18.6% | 16.3% | 10.8% | 9.3% | 16.7% |
| Observations | 778 | 91 | 91 | 91 | 91 | 91 | 91 | 91 |

The distribution of the 5-day average of deaths shows that the growth in deaths is monotonically decreasing over time, but less and less strongly as time passes, supporting the assumption of our linear plus quadratic specification. In week 1 mortality is increasing in all countries, after which it begins to decline for many but not all observations, and finally the downward trend seems to plateau by weeks 6-7.

**Table A2:** Effect of Salience and Closure on Daily Change in Deaths using Fixed Effects

|  |  |  |  |  |
| --- | --- | --- | --- | --- |
|  | Model I | Model II | Model III | Model IV |
| Days from $t_0$ | -0.96 (0.000) | -0.93 (0.000) | -0.99 (0.000) | -0.73 (0.000) |
|  | [-1.2, -0.71] | [-1.2, -0.70] | [-1.2, -0.77] | [-1.01, -0.45] |
| Days from $t_0$ squared | 0.0094 (0.000) | 0.0091 (0.000) | 0.0097 (0.000) | 0.0070 (0.000) |
|  |  | [0.0067, 0.11] | [0.0074, 0.012] | [0.0042, 0.0098] |
| Binary mobility | -9.4 (0.004) | -10.4 (0.001) | -9.2 (0.001) |  |
|  | [-15.0, -3.7] | [-15.9, -4.9] | [-13.7, -4.7] |  |
| Continuous mobility |  |  |  | -11.9 (0.019) |
|  |  |  |  | [-21.5, -2.3] |
| Binary closure | -12.1 (0.000) | -13.6 (0.000) | -10.9 (0.000) |  |
|  | [-16.2, -8.1] | [-17.8, -9.4] | [-13.8, -8.0] |  |
| Continuous closure |  |  |  | -21.1 (0.000) |
|  |  |  |  | [-28.0, -14.3] |
| Country fixed effects? | Yes | Yes | Yes | Yes |
| Number of observations | 778 | 778 | 778 | 776 |
| Notes: p-values are presented in parentheses, 95% confidence intervals are presented in square brackets. Standard errors are clustered at the country level. N lower in Model IV because the lagged mobility data is only available for Italy from the third day of the epidemic. |  |  |  |  |

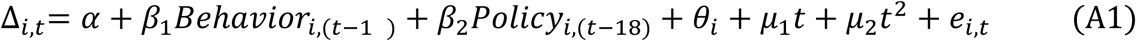

Table A2 presents the results running Models I-IV in Table 2 using a fixed effect specification illustrated in Equation (A1). The model includes a *θ*_*i*_ term to capture country-specific variation of the kind described above, and excludes *X*_*i*_ due to collinearity. However, while it does not require the random effects assumption (that conditional on country controls all other country-specific variation is distributed randomly), the fixed effects model is less efficient compared with the random effects model. We run a Hausman test, without clustering standard errors, to confirm the validity of the random effects assumption by comparing the estimates in our fixed effects model to our random effects model with controls. The results confirm the random effects assumption, and so we use the random effects model as our primary specification.

**Table A3:** Substituting Inherent Salience for Observed Mobility and Equalizing Length of Time

|  |  |  |  |
| --- | --- | --- | --- |
|  | Model I | Model II | Model III |
| Days from $t_0$ | -0.88 (0.000) | -1.1 (0.000) | -1.06 (0.000) |
|  | [-1.1, -0.67] | [-1.3, -0.86] | [-1.6, -0.59] |
| Days from $t_0$ squared | 0.0087 (0.000) | 0.011 (0.000) | 0.012 (0.005) |
|  | [0.0062, 0.011] | [0.0085, 0.013] | [0.0035, 0.020] |
| Binary mobility | -9.2 (0.000) |  | -7.9 (0.001) |
|  | [-14.0, -4.5] |  | [-12.8, -3.1] |
| Inherent salience |  | -2.07 (0.409) |  |
|  |  | [-7.0, 2.8] |  |
| Binary closure | -14.0 (0.000) | -8.3 (0.001) | -12.4 (0.000) |
|  | [-17.2, -10.8] | [-13.3, -3.3] | [-15.3, -9.4] |
| Country fixed effects? | No | No | No |
| Number of observations | 778 | 778 | 650 |
| Notes: Model III limits the number of days of data in each regression to 50, which is the lowest total in the group of countries (Ireland). |  |  |  |

Mobility decline has the advantage of objectively measuring behavior, but it only captures a subset of the relevant aspects – in particular nothing to do with personal hygiene (e.g. handwashing) or with maintaining distance when interacting with others. Although we cannot directly measure these, we can proxy for them by noting that they greatly increase in prevalence when the pandemic becomes locally salient, approximated when the average daily number of deaths reaches 5 in any given country (i.e. at time t_0_; table 1). We refer to this measure as *inherent salience*, switching from 0 to 1 at time t0, as long as that is before binary closure takes effect, and switching off again when closure applies. Model II of Table A2 tests this measure of inherent salience. We find that the coefficient is negative, but insignificant. Closure is still negative and significant, but the effect size is reduced. Separately, in our main specification some countries are effectively more heavily weighted than others since we have more data available for countries whose epidemics began earlier. Model III removes all datapoints later than the shortest epidemic length (Ireland at 50 days). Compared to Model I we find that the coefficients are still significant and roughly the same magnitude, suggesting that this country weighting effect does not introduce a material bias.

## Notes

### Competing Interest Statement

The authors have declared no competing interest.

### Funding Statement

No external funding was received for this research.

